# Epidemics forecast from SIR-modeling, verification and calculated effects of lockdown and lifting of interventions

**DOI:** 10.1101/2020.08.12.20173294

**Authors:** R. Schlickeiser, M. Kröger

## Abstract

Due to the current COVID-19 epidemic plague hitting the worldwide population it is of utmost medical, economical and societal interest to gain reliable predictions on the temporal evolution of the spreading of the infectious diseases in human populations. Of particular interest are the daily rates and cumulative number of new infections, as they are monitored in infected societies, and the influence of non-pharmaceutical interventions due to different lockdown measures as well as their subsequent lifting on these infections. Estimating quantitatively the influence of a later lifting of the interventions on the resulting increase in the case numbers is important to discriminate this increase from the onset of a second wave. The recently discovered new analytical solutions of Susceptible-Infectious-Recovered (SIR) model allow for such forecast and the testing of lockdown and lifting interventions as they hold for arbitrary time dependence of the infection rate. Here we present simple analytical approximations for the rate and cumulative number of new infections.

## Introduction

The Susceptible-Infectious-Recovered (SIR) model has been developed nearly hundred years ago^1,2^ to understand the time evolution of infectious diseases in human populations. The SIR system is the simplest and most fundamental of the compartmental models and its variations.^3–15^ The considered population of *N* ≫ 1 persons is assigned to the three compartments *s* (susceptible), *i* (infectious), or *r* (recovered/removed). Persons from the population may progress with time between these compartments with given infection (*a*(*t*)) and recovery rates (*µ*(*t*)) which in general vary with time due to non-pharmaceutical interventions taken during the pandemic evolution.

Let *I*(*t*) = *i*(*t*)*/N*, *S*(*t*) = *s*(*t*)*/N* and *R*(*t*) = *r*(*t*)*/N* denote the infected, susceptible and recovered/removed fractions of persons involved in the infection at time *t*, with the sum requirement *I*(*t*)+ *S*(*t*)+ *R*(*t*) = 1. In terms of the reduced time 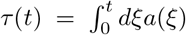, accounting for arbitrary but given time-dependent infection rates, the SIR-model equations are^1,2,16^

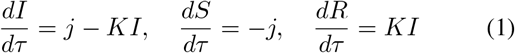

in terms of the time-dependent ratio *K*(*t*) = *µ*(*t*)*/a*(*t*) of the recovery and infection rates and the medically interesting daily rate of new infections

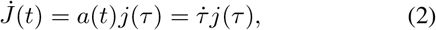

where the dot denotes a derivative with respect to *t*.

For the special and important case of a time-independent ratio *K*(*t*) = *k* = const. new analytical results of the SIR-model (1) have been recently derived^17^ – hereafter referred to as paper A. The constant *k* is referred to as the inverse basic reproduction number *k* = 1*/R*_0_. The new analytical solutions assume that the SIR equations are valid for all times *t* ∈ [−∞, ∞], and that time *t* = *τ =* 0 refers to the ‘observing time’ when the existence of a pandemic wave in the society is realized and the monitoring of newly infected persons 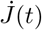 is started. In paper A it has been shown that, for arbitrary but given infection rates *a*(*t*), apart from the peak reduced time *τ*_0_ of the rate of new infections, all properties of the pandemic wave as functions of the reduced time are solely controlled by the inverse basic reproduction number *k*. The dimensionless peak time *τ*_0_ is controlled by *k* and the value *ε* = − ln *S*(0), indicating as only initial condition at the observing time the fraction of initially susceptible persons *S*(0) = *e^−ε^*. This suggests to introduce the relative reduced time Δ = *τ* − *τ*_0_ with respect to the reduced peak time. In real time *t* the adopted infection rate *a*(*t*) acts as second parameter, and the peak time *t_m_*, where 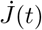 reaches its maximum must not coincide with the time, where the reduced *j* reaches its maximum, i.e., *τ_m_* ≡ *τ*(*t_m_*) = *τ*_0_, in general. *SIR-model results:* According to paper A the three fractions of the SIR-model

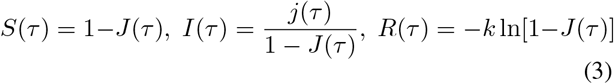

can be expressed in terms of the cumulative number *J*(*τ*) and differential daily rate *j*(*τ*) = *dJ/dτ* of new infections. The cumulative number satisfies the nonlinear differential equation

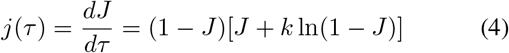

Two important values are *J*_0_(*k*) = *J*(*τ*_0_), where *j* attains its maximum with 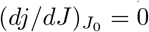, and the final cumulative number *J*_∞_(*k*) at *τ* = *t* = ∞, when the second bracket on the right-hand side of the differential equation (4) vanishes, i.e. *J*_∞_ + *k* ln(1 − *J*_∞_) = 0. The two transcendental equations can be solved analytically in terms of Lambert’s *W* function, as shown in paper A. In the present manuscript we are going to avoid Lambert’s function completely, and instead use the following approximants (Fig. 1a)

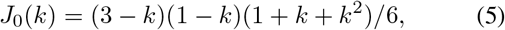

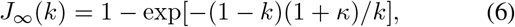

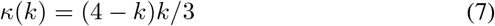

Without any detailed solution of the SIR-model equations the formal structure of Eqs. (3) and (4) then provides the final values *I*_∞_ = *j*_∞_ = 0, *R*_∞_ = *J*_∞_, and *S*_∞_ = 1 − *J*_∞_. We list these values together with *κ* in Tab. I.

**FIG. 1.**
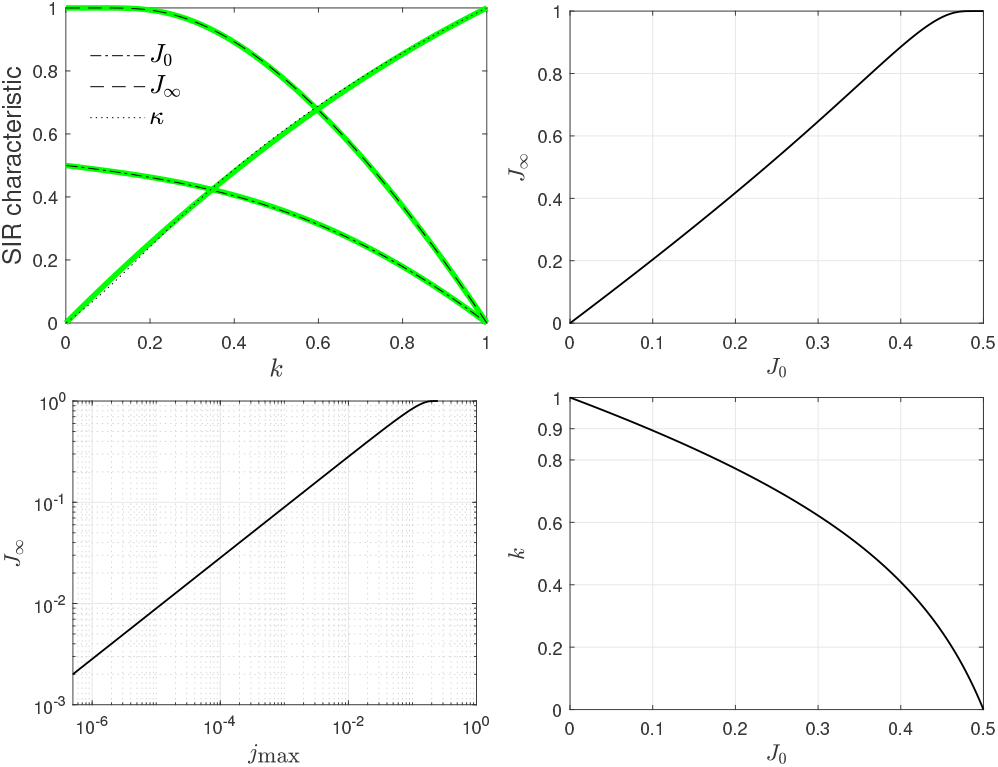
**(a)** Approximants *J*_0_, *J*_∞_, and *κ* (thick green) used in this manuscript, cf. Eqs. (5)–(7), compared with the exact functions^17^ (thin black). **(b)** *J*_∞_ versus *J*_0_ and **(c)** *J*_∞_ versus *j*_max_ for the SIR model. **(d)** *k* as function of *J*_0_ according to Eq. (14). For *J*_0_ ≤ 0.1 this is well approximated by *k* ≈ 1 − *J*_0_, where *J*_0_ can be replaced by the cumulative fraction of infected people at the time of the maximum in the daily number of newly infected people.

New infections:

The exact solution of the differential equation (4) is given in inverse form by (Appendix A)

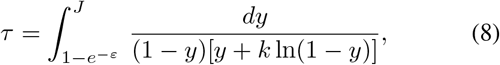

which can be integrated numerically (subject to numerical precision issues), replaced by the approximant presented in paper A (involving Lambert’s function), or semi-quantitatively captured by the simple approximant to be presented next. The solution *J*(Δ) as a function of the relative reduced time Δ = *τ* − *τ*_0_, with the reduced peak time approximated by

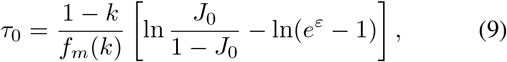

corresponding to *J* = *J*_0_ in Eq. (8), and where *f_m_*(*k*) = 1 − *k* + ln *k*, is reasonably well captured by (Appendix C)

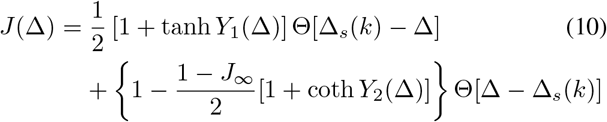

with the Heaviside step function *Θ*(*x*) = 1(0) for *x* ≥ (*<*)0. In Eq. (10)

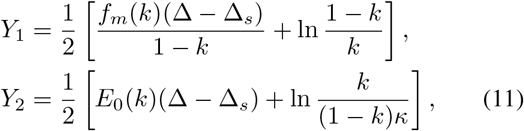

with

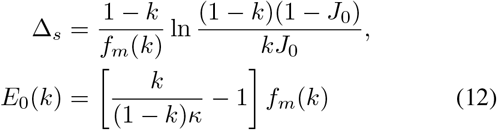

also tabulated in Tab. I. We note that Δ*_s_*(*k*) is always positive. Figure 2 shows the approximation (10) for the cumulative number as a function of the relative reduced time Δ for different values of *k*. For a comparison with the exact variation obtained by the numerical integration of Eq. (8) see Appendix C. The agreement is remarkably well with maximum deviations less than 30 percent.

**FIG. 2.**
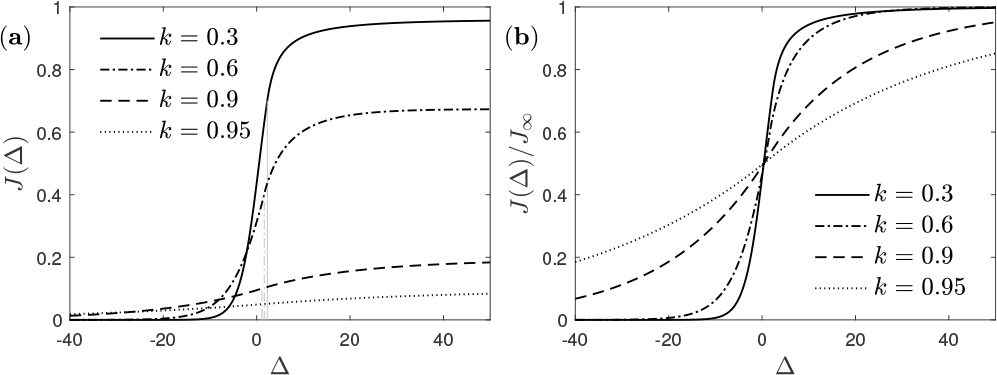
**(a)** Cumulative number *J*(Δ) for different values of *k* according to Eq. (10). The vertical gray lines starting at the Δ-axis indicate the respective values of Δs(*k*).**(b)** Same as in (a), divided by the final *J*_∞_.

**Table I.** Exact parameter values depending on the inverse basic reproduction number *k*.

| $k$ | $\kappa$ | $J_0$ | $J_\infty$ | $S_\infty$ | $f_m$ | $E_0$ | $\Delta_s$ | $j_{\max}$ |
| --- | --- | --- | --- | --- | --- | --- | --- | --- |
| 0.00 | 0.00 | -Inf | 1.000 | 0.000 | 1.000 | 0.0000 | ∞ | 0.2500 |
| 0.05 | 0.05 | 0.492 | 1.000 | 0.000 | 0.800 | 0.0000 | 3.5339 | 0.2327 |
| 0.10 | 0.11 | 0.483 | 1.000 | 0.000 | 0.670 | 0.0003 | 3.0441 | 0.2156 |
| 0.15 | 0.17 | 0.473 | 0.999 | 0.001 | 0.565 | 0.0049 | 2.7698 | 0.1986 |
| 0.20 | 0.24 | 0.462 | 0.993 | 0.007 | 0.478 | 0.0173 | 2.5745 | 0.1819 |
| 0.25 | 0.31 | 0.450 | 0.980 | 0.020 | 0.403 | 0.0348 | 2.4181 | 0.1653 |
| 0.30 | 0.37 | 0.436 | 0.959 | 0.041 | 0.339 | 0.0535 | 2.2835 | 0.1490 |
| 0.35 | 0.43 | 0.421 | 0.930 | 0.070 | 0.283 | 0.0709 | 2.1621 | 0.1330 |
| 0.40 | 0.49 | 0.403 | 0.893 | 0.107 | 0.234 | 0.0857 | 2.0491 | 0.1174 |
| 0.45 | 0.54 | 0.384 | 0.848 | 0.152 | 0.191 | 0.0971 | 1.9418 | 0.1022 |
| 0.50 | 0.59 | 0.363 | 0.796 | 0.203 | 0.153 | 0.1050 | 1.8386 | 0.0876 |
| 0.55 | 0.64 | 0.339 | 0.739 | 0.261 | 0.121 | 0.1093 | 1.7386 | 0.0736 |
| 0.60 | 0.69 | 0.313 | 0.676 | 0.324 | 0.094 | 0.1099 | 1.6416 | 0.0603 |
| 0.65 | 0.73 | 0.283 | 0.607 | 0.393 | 0.070 | 0.1071 | 1.5477 | 0.0479 |
| 0.70 | 0.78 | 0.251 | 0.533 | 0.467 | 0.050 | 0.1009 | 1.4571 | 0.0365 |
| 0.75 | 0.82 | 0.217 | 0.454 | 0.546 | 0.034 | 0.0914 | 1.3701 | 0.0263 |
| 0.80 | 0.86 | 0.179 | 0.371 | 0.629 | 0.022 | 0.0788 | 1.2871 | 0.0174 |
| 0.85 | 0.89 | 0.138 | 0.284 | 0.716 | 0.012 | 0.0633 | 1.2084 | 0.0102 |
| 0.90 | 0.93 | 0.095 | 0.193 | 0.807 | 0.005 | 0.0449 | 1.1343 | 0.0047 |
| 0.95 | 0.97 | 0.049 | 0.098 | 0.902 | 0.001 | 0.0237 | 1.0648 | 0.0012 |
| 1.00 | 1.00 | 0.000 | 0.000 | 1.000 | 0.000 | 0.0000 | 1.0000 | 0.0000 |

For the corresponding reduced differential rate *j*(Δ) in reduced time we use the right hand side of Eq. (4) with *J* = *J*(Δ) from Eq. (10), cf. Fig. 3. Note, that this *j* is not identical with the one obtained via *j* = *dJ/dτ*, because *J* does not solve the SIR equations exactly. The peak value *j*_max_ in the reduced time rate occurs when *J* = *J*_0_ and is thus determined by *j*_max_ = (1 − *J*_0_)(1 − *J*_0_ − *k*), also tabulated in Tab. I.

**FIG. 3.**
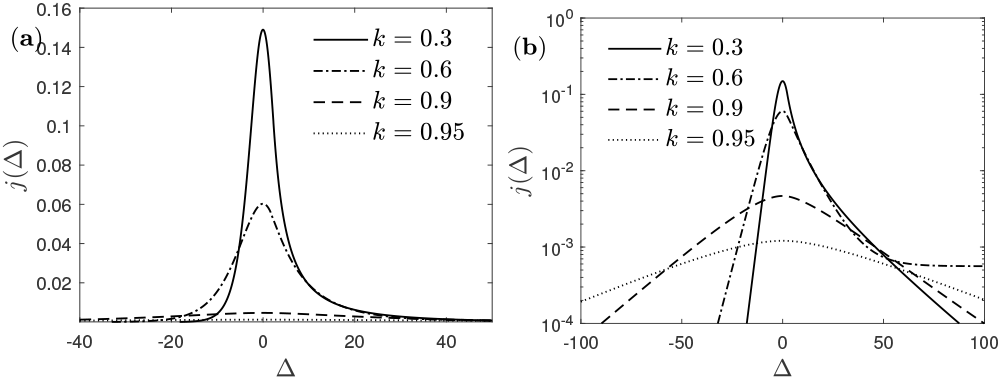
Reduced differential rate *j*(Δ) of newly infected fraction corresponding to the cumulative *J*(Δ) shown in Fig. 2. **(a)** linear scale, **(b)** semilogarithmic scale.

As can be seen in Fig. 3 the rate of new infections (12) is strictly monoexponentially increasing 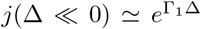 with *Γ*_1_(*k*) = *f_m_*(*k*)*/*(1 − *k*) well before the peak time, and strictly monoexponentially decreasing well above the peak time 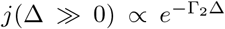 with the *Γ*_2_ = (1 − *J*_∞_)*Γ*_1_*/κ*. These exponential rates exhibit a noteworthy property and correlation in reduced time:

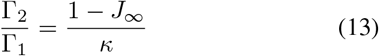

The SIR parameter *k* affects several key properties of the differential and cumulative fractions of infected persons. If the maximum 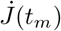 of the measured daily number of newly infected persons has passed already, we find it most convenient to estimate *k* from the cumulative value *J*(*t_m_*) at this time *t_m_*. While the maximum of 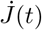 must not occur exactly at *τ* (*t_m_*) = *τ*_0_, we can still use *J*_0_ as an approximant for the value of *J*(*t_m_*) and the relationship between *J*_0_ and *k* can be inverted to read (Appendix E)

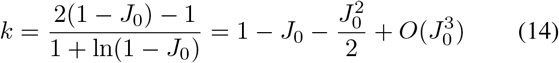

The dependency of *k* on *J*_0_ is shown in Fig. 1c. With the so-obtained value for *k* at hand, the infection rate *a*(*t_m_*) at peak time can be inferred from 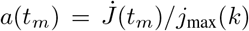. It provides a lower bound for *a*_0_.

A major advantage of the new analytical solutions in paper A and here is their generality in allowing for arbitrary time-dependencies of the infection rate *a*(*t*). Such time-dependencies result at times greater than the observing time *t* = 0 from non-pharmaceutical interventions (NPIs) taken after the pandemic outbreak^18^ such as case isolation in home, voluntary home quarantine, social distancing, closure of schools and universities and travel restrictions including closure of country borders, applied in different combinations and rigour^19^ in many countries. These NPIs lead to a significant reduction of the initial constant infection rate *a*_0_ at later times. It is also important to estimate the influence of a later lifting of the NPIs on the resulting increase in the case numbers in orderr to discriminate this incresase from the onset of a second wave.

## Modeling in real time of lockdowns

The corresponding daily rate 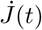 and cumulative number *J*(*t*) of new infections in real time *t* for given time-dependent infection rates *a*(*t*) are *J*(*t*) = *J*(*τ* (*t*)) and 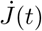 given by Eq. (2). From a medical point of view the daily rate 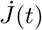 is most important as it determines also (i) the fatality rate^20^ 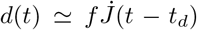 with the fatality percentage *f* ⋍ 0.005 in countries with optimal medical services and hospital capacities and the delay time of *t_d_* ⋍ days, (ii) the daily number of new seriously sick persons^21^ NSSPs = 2*f* 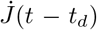 needing access to breathing apparati, and (iii) the day of maximum rush to hospitals *t_r_* = *t_m_* + *t_d_*. In countries with poor medical and hospital capacities and/or limited access to them the fatality percentage is significantly higher by a factor *h* which can be as large as 10.

To calculate the rate and cumulative number in real time according to Eq. (2) we adopt as time-dependent infection rate the integrable function known from shock wave physics

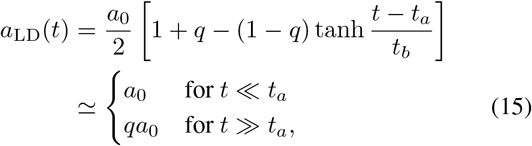

which implies

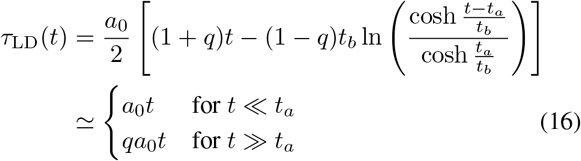

The time-dependent lockdown infection rate (15) is characterized by four parameters: (i) the initial constant infection rate *a*_0_ at early times *t* ≪ *t_a_*, (ii) the final constant infection rate *a*_1_ = *qa*_0_ at late times *t* ≫ *t_a_* described by the quarantine factor *q* = *a*_1_*/a*_0_ ≤ 1, first introduced in Refs.^19,21^, (iii) the time *t_a_* of maximum change, and (iv) the time *t_b_* regularizing the sharpness of the transition. The latter is known to be about *t_b_* 7–14 days reflecting the typical one-two weeks incubation delay.

Moreover, the initial constant infection rate *a*_0_ characterizes the Covid-19 virus: if we adopt the German values *a*_0_ ⋍ 58 days^−1^ and *t_b_* ⋍ 11 determined below, with the remaining two parameters *q* and *t_a_* we can represent with the chosen functional form (15) four basic types of reductions: (1) drastic (small *q* « 1) and rapid (*t_a_* small), (2) drastic (small *q* ≪ 1) and late (*t_a_* large), (3) mild (greater *q*) and rapid (*t_a_* small), and (4) mild (greater *q*) and late (*t_a_* large). The four types are exemplified in Fig. 4.

**FIG. 4.**
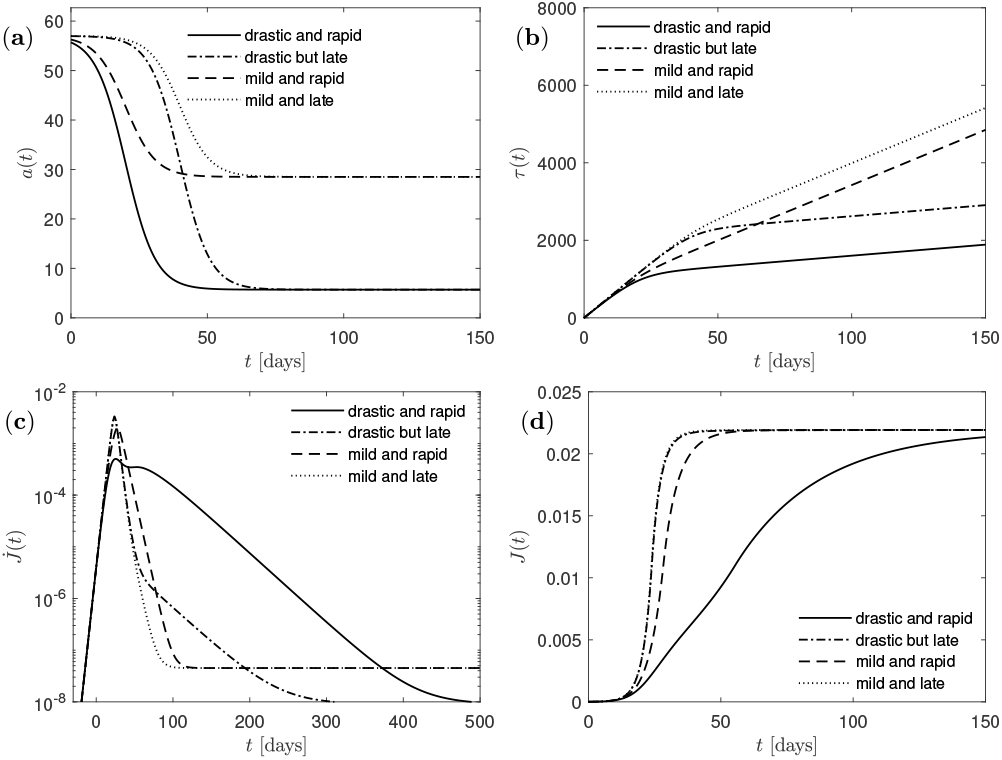
**(a)** infection rate *a*(*t*),**(b)** reduced time *τ* (*t*),**(c)** daily rate of new infections 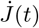, **(d)** cumulative fraction *J*(*t*) of infected persons. In each panel we consider four basic types of reductions: (1) drastic (small *q* = 0.1) and rapid (*t*a = 20), (2) drastic (small *q* = 0.1) and late (*t*a = 40), (3) mild (*q* = 0.5) and rapid (*t*a = 20), and (4) mild (*q* = 0.5) and late (*t*a = 40). Remaining parameters due to Germany: *t*b = 11 days, *a*0 = 57 days_−_^1^, and *k* = 0.989.

## Verification and forecast

In countries where the peak of the first Covid-19 wave has already passed such as e.g. Germany, Switzerland, Austria, Spain, France and Italy, we may use the monitored fatality rates and peak times to check on the validity of the SIR model with the determined free parameters. However, later monitored data are influenced by a time varying infection rate *a*(*t*) resulting from non-pharmaceutical interventions (NPIs) taken during the pandemic evolution. Only at the beginning of the pandemic wave it is justified to adopt a time-independent injection rate *a*(*t*) *a*_0_ implying *τ* = *a*_0_*t*. Alternatively, also useful for other countries which still face the climax of the pandemic wave, it is possible to determine the free parameters from the monitored cases in the early phase of the pandemic wave. We illustrate our parameter estimation using the monitored data from Germany with a total population of 83 million persons (*P* = 8.3 × 10^7^).

In Germany the first two deaths were reported on March 9 so that *ε* = 4.8 × 10^−6^ corresponding to about 400 infected people 7 days earlier, on March 2 (*t* = 0). The maximum rate of newly infected fraction, 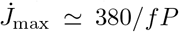, occurred *t_m_* = 37 days later, consistent with a peak time of fatalities on 16 April 2020. At peak time the cumulative death number was *D_m_* = 3820*/P* corresponding to *J_m_* = *D_m_/f* = 200*D*_0_ = 0.009. This implies *k ≈* 1 − *J*_0_ = 0.991 according to Eq. (14). From the initial exponential increase of daily fatalities in Germany we extract Γ_1_(*k*)*a*_0_ ⋍ 0.28, corresponding to a doubling time of ln(2)*/*Γ_1_*a*_0_ 2.3 days, as we know Γ_1_(*k*) = *f_m_*(*k*)*/*(1 − *k*)0.0046 already from the above *k*. The quantity *a_m_* we can estimate from the measured 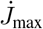, as 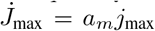 and *j*_max_(*k*) ≈ 4.2 × 10^−5^. Using the mentioned value for 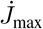, we obtain *a_m_* ≈ 22*/*days as a lower bound for *a*_0_.

With these parameter values the entire following temporal evolution of the pandemic wave in Germany can be predicted as function of *t_b_* and *q*. To obtain all parameters consistently, we fitted the available data to our model without constraining any of the parameters (Fig. 5). This yields for Germany *k* ⋍ 0.989, *t_a_* ⋍ 21 days, *q* ⋍ 0.15, *a*_0_ ⋍ 0 58 days, and *t_b_* = 11 days. The obtained parameters allow us to calculate the dimensionless peak time *τ_m_* 1353, the dimensionless time *τ*_0_ ⋍ 1390, as well as *J_m_* ⋍ 0.009, *J*_0_ ⋍ 0.011, Γ_1_ ⋍ 0.0056 and Γ_2_ ⋍ 0.0055.

**FIG. 5.**
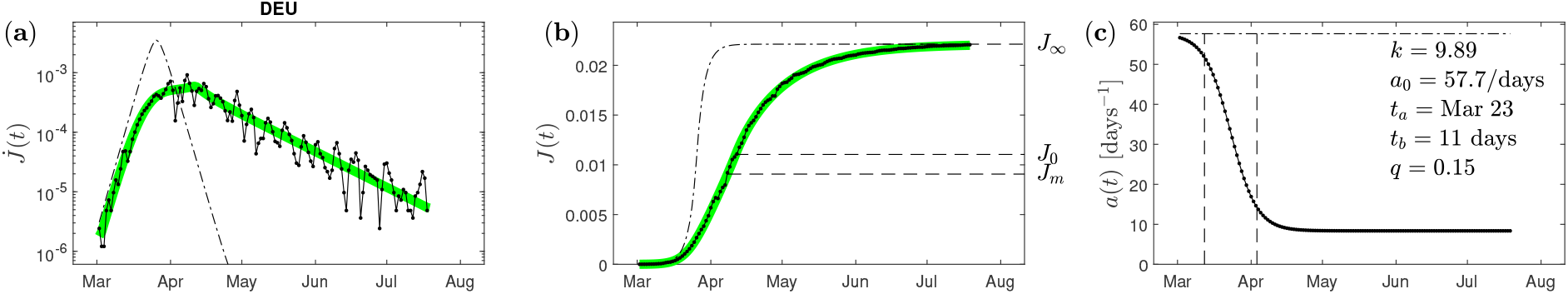
**(a)** Measured data 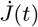 of new daily infected fraction (black circles) for Germany (DEU) compared with the model 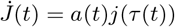 outlined here (green). Shown for comparison is the case where no NPIs are imposed (*q* = 1, black dot-dashed). **(b)** The measured cumulative fraction *J*(*t*) (black circles) together with the model prediction (green), and the reference *q* = 1 case. Also depicted are *J*_0_ and the value at peak time, *J_m_*.**(c)** The infection rate *a*(*t*) corresponding to the curves shown in (a) and (b). Model parameters mentioned in the figure; *a* dropped from an initial value of 58/days down to 7.8/days during the 2nd half of march. This realized case can be directly compared with the four hypothetic cases shown in Fig. 4.

We note that the value of *k* = 0.989 implies for Germany that *J*_∞_(0.989) = 0.022 according to Fig. 1, so that at the end of the first Covid-19 wave in Germany 4 percent of the population, i.e. 1.83 million persons will be infected. This number corresponds to a final fatality number of *D*_∞_ = 9310 persons in Germany. Of course, these numbers are only valid estimates if no efficient vaccination against Covid-19 will be available.

An important consequence of the small quarantine factor *q* = 0.15 is the implied flat exponential decay after the peak. Because Γ_1_ ⋍ Γ_2_ for *k* = 0.989, the exponential decay is by a factor *q* smaller than the exponential rise prior the climax, i.e.,

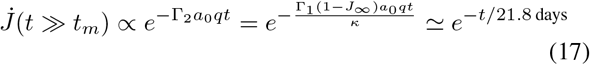

Equation (17) yields a decay half-live of ln(2) × 21.8 days ⋍ 15 days to be compared with the initial doubling time of ⋍ 2.3 days. For Germany we thus know that their lock-down was drastic and rapid: the time *t_a_* March 23 is early compared to the peak time *t_m_* ⋍ Apr 8 resulting in a significant decrease of the infection rate with the quarantine factor *q* ⋍ *a_m_/a*_0_ = 0.15. In Fig. 5 we calculate the resulting daily new infection rate as a function of the time *t* for the parameters for Germany, and compare with the measured data. In the meantime, the strict lockdown interventions have been lifted in Germany: This does not effect the total numbers *J*_∞_ and *D*_∞_ but it should reduce the half-live decay time further.

We also performed this parameter estimation for other countries with sufficient data. For some of them data is visualized in Fig. 7, parameters for the remaining countries are tabulated in Appendix I.

The presented equations hold for arbitrary *a*(*t*). An example, on how lockdown lifting can be modeled is described in Appendix H. The situation is depicted in Fig. 6. The lifting will increase *a*(*t*) from its present value up to a value that might be close to the initial *a*_0_. While the dynamics is altered, the final values remain unaffected by the dynamics, except, if the first pandemic wave is followed by a 2nd one. The values for *J*_∞_ provided in the tables of Appendix I provide a hint on how likely is a 2nd wave. These values correspond to the population fraction that had been infected already. While this fraction is extremely large in Peru (53%), it is still below 1% in several of the larger countries. The tables also report the unreported number of infections per reported number (column ‘dark’), estimated from the number of fatalities, reported infections, and the death probability *f*.

**FIG. 6.**
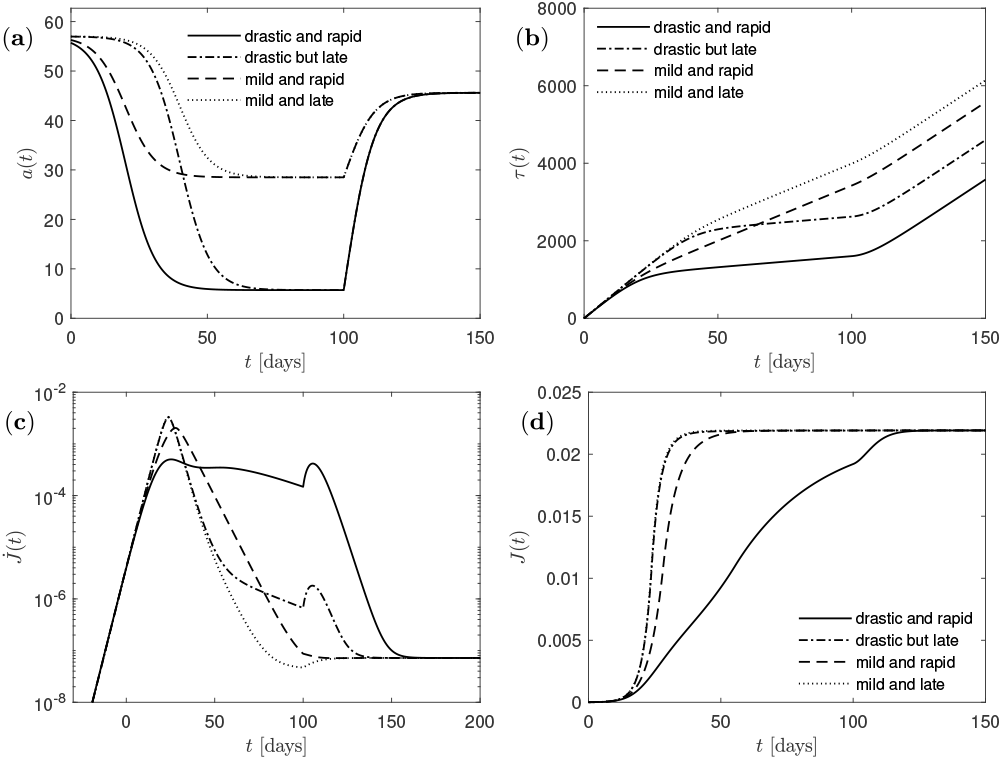
Same as Fig. 4 but with incomplete lifting (*η* = 0.8) hundred days after breakout (*t_s_* = 100 days). **(a)** infection rate *a*(*t*),**(b)** reduced time *τ* (*t*),**(c)** daily rate of new infections 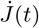, **(d)** cumulative fraction *J*(*t*) of infected persons. In each panel we consider four basic types of reductions: (1) drastic (small *q* = 0.1) and rapid (*t_a_* = 20), (2) drastic (small *q* = 0.1) and late (*t_a_* = 40), (3) mild (*q* = 0.5) and rapid (*t_a_* = 20), and (4) mild (*q* = 0.5) and late (*t_a_* = 40). Remaining parameters due to Germany: *t_b_* = 11 days, *a*_0_ = 57 days^−1^, and *k* = 0.989.

**FIG. 7.**
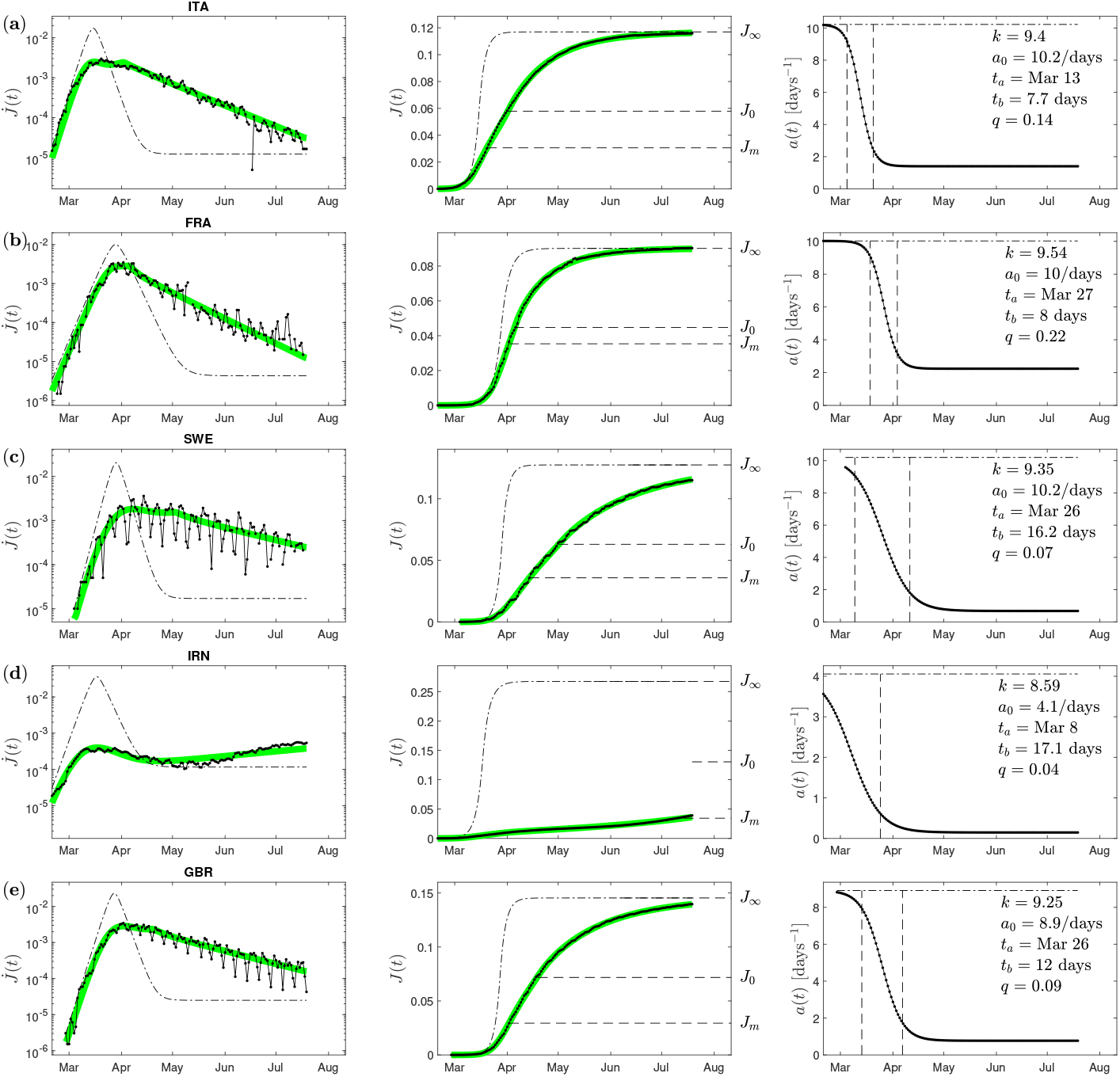
(Same as Fig. 5 for other countries: **(a)** Italy (ITA), **(b)** France (FRA), **(c)** Sweden (SWE), **(d)** Iran (IRN), **(e)** Great Britain (GBR).

## Data Availability

All data is public available from the link provided below.

https://pomber.github.io/covid19/timeseries.json

## Appendix A: Non-parametric solution of the SIR model

We start from the Eq. (A-29) [the notation (A-x) refers to Eq. (x) in paper A]

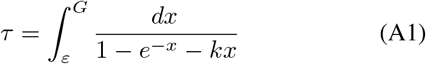

and substitute

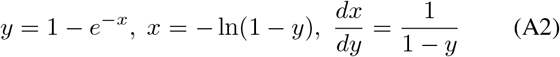

Consequently, as the cumulative number of new infections is given (see Eq. (A-37)) by

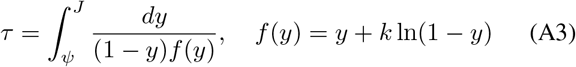

with the abbreviation *ψ* = *J*(0) = 1 − *e^−ε^* for the initial value. This inverse relation *τ*(*J*) is the general solution of the SIR-model for constant *k*. It is not in parametrized form.

### 1. Maximum of *j*

Taking the derivative of Eq. (A3) with respect to *τ* we obtain

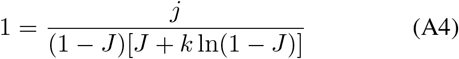

or the exact SIR relation

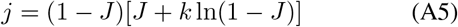

Equation (A5) provides

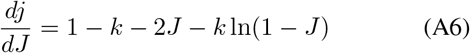

The maximum value *j*_max_ occurs for 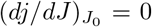 providing

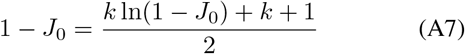

Setting 1 − *J*_0_ = *e*^−^*^X^* yields

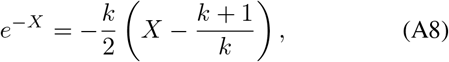

which is of the form (A-G1), and solved in terms of the non-principal Lambert function *W*_−1_ as>

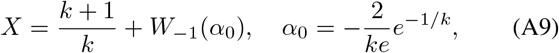

so that

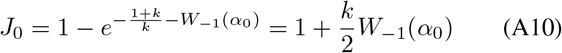

The maximum value is then given by

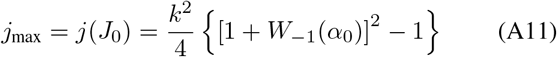

and this can also be written as *j*_max_ = (1 − *J*_0_)(1 − *J*_0_ − *k*) with *J*_0_ from Eq. (A10). According to Eq. (4) the reduced peak time in the dimensionless rate of new infections is then given by

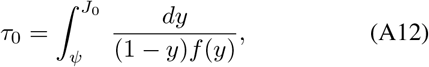

which is the only quantity depending besides on *k* also on *ε* via *ψ* = 1 − *e*^−^*^ε^*. In order to have one approximation depending only on *k* we therefore introduce the relative reduced time with respect to the peak reduced time

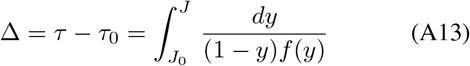

Equation (4) then becomes

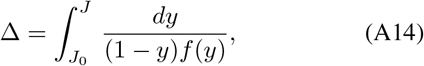

which is still exact, independent of *ε* and only determined by the value of *k*.

## Appendix B: Approximating the function *f*(*y*)

The function *f*(*y*) defined in Eq. (A3) vanishes for 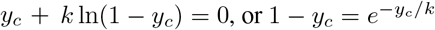 with the solution

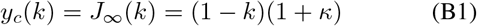

where *κ* was already stated in the introduction. According to Eq. (A14) the value *J*_∞_ corresponds to Δ = *τ* = ∞, so the maximum value of the cumulative number of new infections is *J*_max_ = *J*_∞_.

Moreover, the function *f*(*y*) attains its maximum value

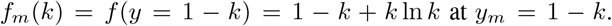

As approximation we use

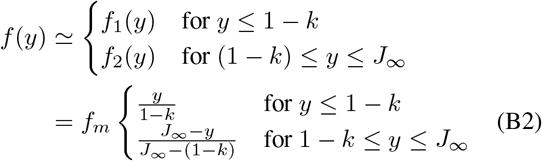

which is shown in Fig. 8 in comparison with the function *f*(*y*). The agreement is reasonably well with maximum deviations less than 30 percent.

**FIG. 8.**
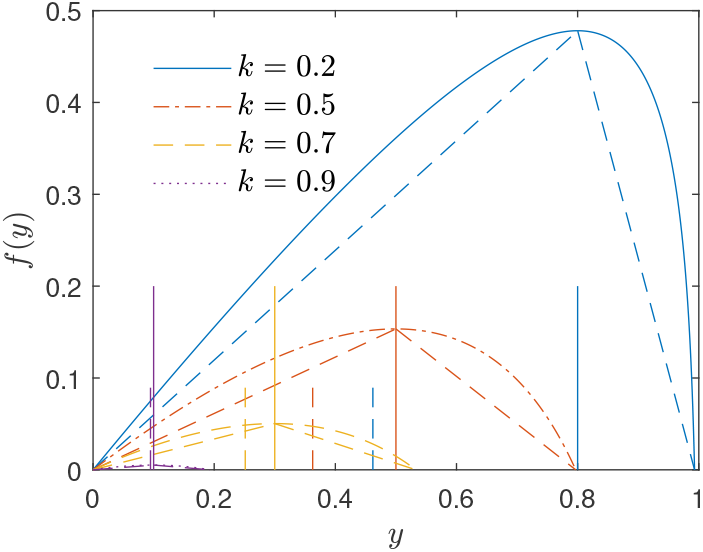
Comparison of the approximation (B2) with the exact curve for *f*(*y*) for different values of *k*.

## Appendix C: Approximations for *J*(*τ*)

Figure 9 demonstrates that *J*_0_(*k*) is always smaller than 1 − *k*. In order to calculate the integral in Eq. (A14) with the approximation (B2) we then have to investigate two cases: (1) For *J*_0_ *<* 1−*k* and *J<* 1−*k* only the function *f*_1_ contributes and

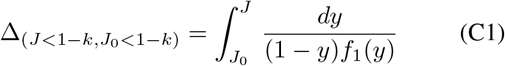

**FIG. 9.**
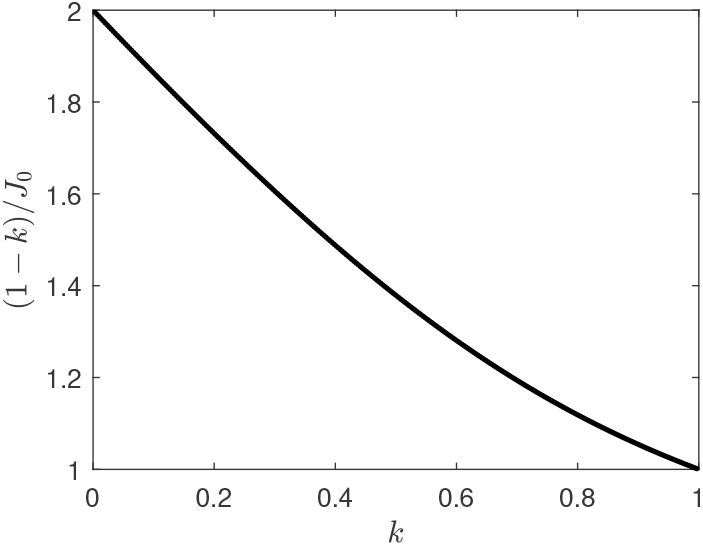
The ratio (1 − *k*)*/J*_0_(*k*) as a function of *k*.

(2) For *J ≥* 1 − *k>J*_0_ both functions *f*_1_ and *f*_2_ contribute and

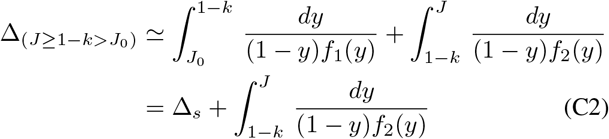

with

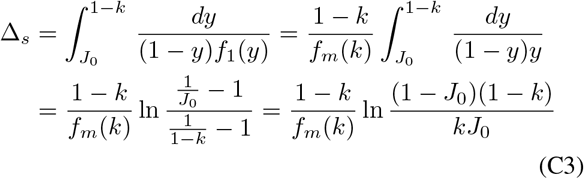

(C3) denoting the relative time corresponding to the value *J* = 1 − *k*. We consider each case in turn.

### 1. **Case (1):** *J* ≤ 1 − *k*, *J*_0_ *<* 1 − *k*

Here Eq. (C1) provides

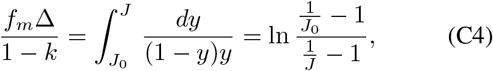

so that the difference of Eqs. (C3) and (C4) yields

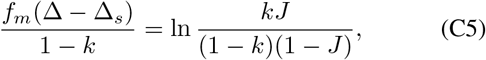

or after inversion

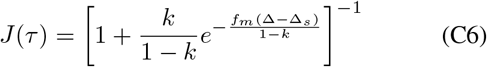

### 2. **Case (2):** *J* ≥ 1 − *k>J*0

Here Eq. (C2) with Eq. (C3) yields

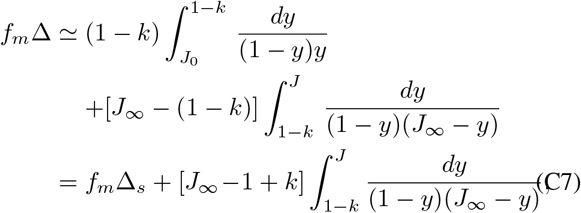

so that

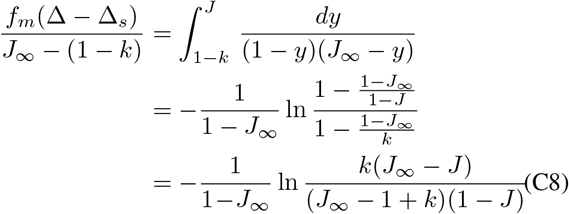

After straightforward but tedious algebra we obtain

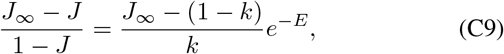

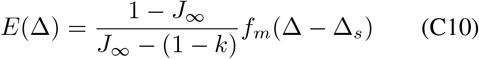

and consequently

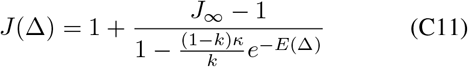

Using the identities 2(1 + *e*^−2^*^Y^*)^−1^ = 1 + tanh *Y* and 2(1 − *e*^−2^*^Y^*)^−1^ = 1 + coth *Y* we combine the results (C6) and (C11) to the analytical approximation of the SIR-model equations at all reduced times, stated in Eqs. (10), (11), and (12). A comparison with the exact numerical solution of the SIR model is provide in Fig. 10. The corresponding *j*(Δ) is obtained from *J*(Δ) via Eq. (A5).

**FIG. 10.**
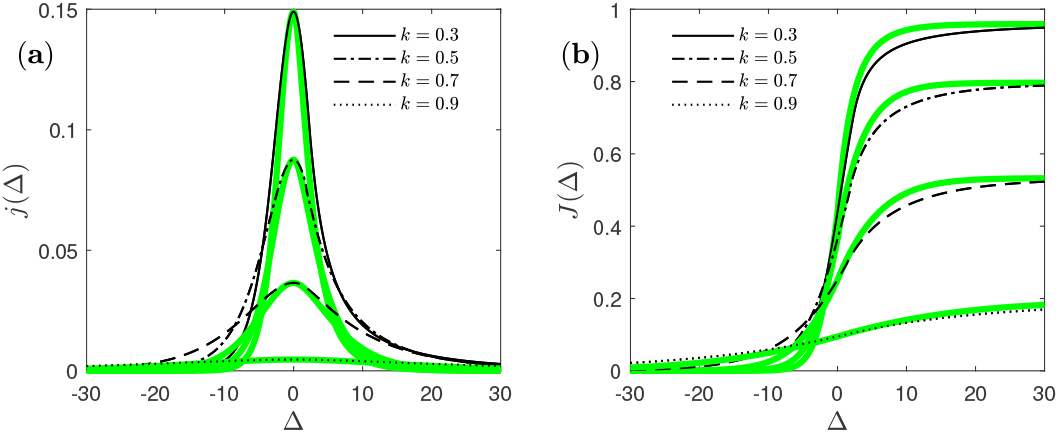
Comparison for **(a)** *j*(Δ) and **(b)** *J*(Δ) between exact solution of the SIR model (green) and the approximant used here (black) for various *k*. Our approximant^22^ in terms of Lambert’s function is shown as well, but coincides with the exact solution (black) in this representation.

## Appendix D: SI-limit *k* = 0

In the limit *k* = 0 Eq. (A7) provides *J*_0_(*k* = 0) = 1*/*2 so that with lim*_k_*_→0_ *f_m_*(*k* = 0) = 1 the time scale (C3) becomes

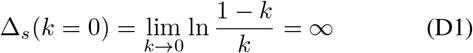

With this result

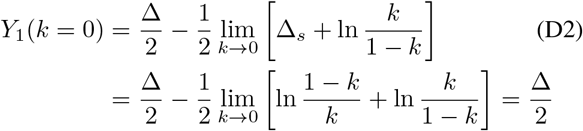

Consequently, the cumulative number (10) and the rate (4) in this case for all times correctly reduce to

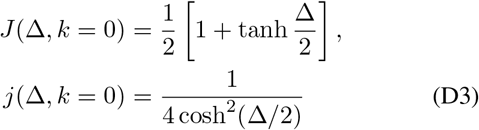

## Appendix E: Relationship between *J*_0_ and *k*

Here we prove Eq. (14). According to paper A the quantity *J*_0_ is given by 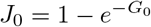 with

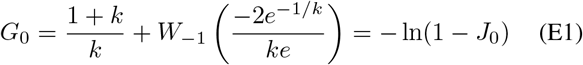

where *e* denotes Euler’s number and *W*_−1_ the non-principal solution of Lambert’s equation *z* = *We^W^*. Equation (E1) is of the form *x* = *r* + *c*^−1^*W* (*ce*^−^*^cr^/β*) upon identifying *c* = 1, *r* = 1*/k*, *β* = −*ke/*2, and *x* = −[1+ln(1 − *J*_0_)]. From paper A we thus know that *e^−cx^* = *β*(*x* − *r*) holds, or equivalently

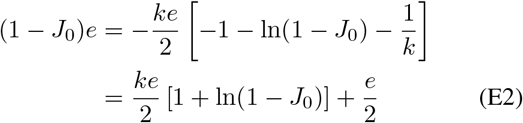

This is readily solved for *k*, and thus proves Eq. (14).

## Appendix F: Time of maximum in the measured differential rate 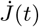

One has *J*(*t*) = *J*(*τ*(*t*)) and 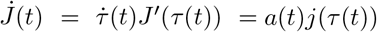 since *j* = *J′* if we let the prime denote a derivative with respect to *τ*. The maximum in 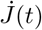 thus fulfills

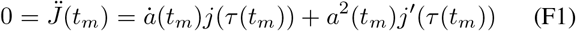

or equivalently,

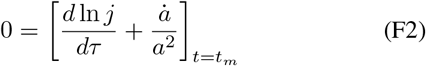

From part A we know that

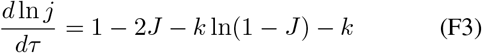

and our *J*_0_ = *J*(*τ*_0_) solves 1 = 2*J*_0_ + *k* ln(1 − *J*_0_)+ *k*. That is, *j*′(*τ*_0_) = 0. If *a* does not depend on time, *τ*_0_ = *τ*(*t_m_*) = *a*_0_*t_m_*, but this is not generally the case. To find *t_m_* and *τ_m_* ≡ *τ* (*t_m_*) one has to solve Eq. (F1), or Eq. (F2). Eq. (F2) with (F3) is solved by

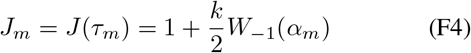

with

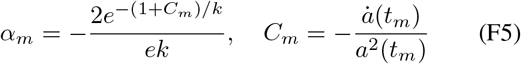

The corresponding *j* is, according to Eq. (4),

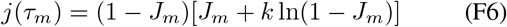

The smaller *C_m_*, the closer is *J_m_* to *J*_0_.

## Appendix G: Fitting the data

As discussed in length in paper A we base our analysis of existing data on the reported cumulative number of deaths, *D*(*t*), from which we estimate the cumulative number of infections *J*(*t*) = *D*(*t* − *t_d_*)*/f* = 200*D*(*t* − *t_d_*) with *t_d_* = 7 days. From the cumulative value *J_m_* = *J*(*t_m_*) at the time *t_m_* of the maximum in 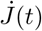 we estimate *k* via Eq. (14) upon assuming *J_m_* ≈ *J*_0_. Similarly, *a_m_* = *a*(*t_m_*) is estimated from 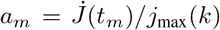. These *t_m_*, *k*, *a_m_* are not the final values, but provide starting values which are then used in the minimization of the deviation between measured and modeled *J*(*t*). The minimization is performed assuming the time-dependent *a*(*t*) parameterized by Eq. (H1) involving parameters *t_a_ >* 0, *t_b_ >* 0, *a*0 *>* 0, *q* ∈ [0, 1], *q<η* ∈ [0, 1] and *t_s_ >t_m_*. While *τ*(*t*) is given by the integrated *a*(*t*), we use three strategies to model *J*(*t*): (i) the numerical solution of the SIR model, (ii) The approximant *G*(*τ*) and *J*(*τ*) = 1−*e*developed in part A, and (iii) the approximant *J*(Δ) given by Eq. (10) with Δ = *τ* − *τ_m_* and *τ_m_* specified by Eq. (9). Because the numerical solution (i) is extremely well approximated by (ii), and (ii) and (iii) compared to (i) not prone to numerical instabilities at small and large Δ, we present results only for method (iii), as they can be readily reproduced by a reader without Lambert’s function at hand.

## Appendix H: Modeling of lockdown lifting

Similarly to the lockdown modeling a later lifting of the NPIs can be modeled by adopting the infection rate

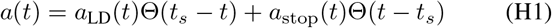

where *t_s_* denotes the stop time of the lockdown still represented by the infection rate (15), and where *a_LD_* is given by Eq. (15). The infection rate after *t_s_* is assumed to be

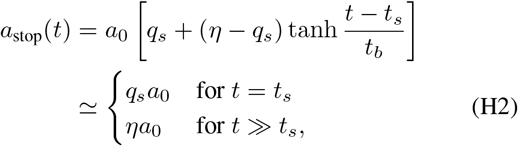

with *q_s_* = *a_LD_*(*t_s_*)*/a*_0_ the quarantine factor reached at the time *t_s_* of lifting. Together with the reduced time (15) we now find

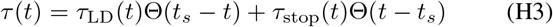

and

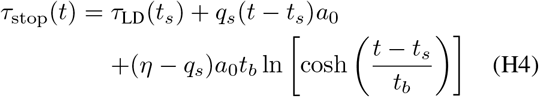

with *τ_LD_*(*t*) from Eq. (16). For the four basic types of Fig. 4 we demonstrate in Fig. 6 the effect of incomplete lifting.

## Appendix I: Analysis for various countries

We performed the parameter estimation for additional countries with sufficient data on 7th Aug 2020. Tables II-III we report these results.

**Table II.** Model parameters and model implications. The columns are as follows: country (*α*_3_ code), population *P* in millions (M), outbreak time defining *t* = 0, fitted time *t_a_*, estimated time *t*_0_ corresponding to the reduced peak time *τ*_0_ of *j*(*τ*), fitted SIR parameter *k*, fitted initial infection rate *a*_0_, fitted parameter *t_b_*, fitted quarantine factor *q*, estimated doubling time *t*_2_ characterizing the early exponential increase, estimated decay half life *t′*_2_ characterizing the late exponential decrease, estimated unreported number of infections per reported number, estimated final fraction *J*_∞_ of infected population, final number of estimated fatalities *D*_∞_*P* = *J*_∞_*Pf*. We use *f* = 0.005 as the probability to decease from a Covid-19 infection (reported plus unreported).

| country | $P/M$ | $t = 0$ | $t_a$ | $t_m$ | $t_0$ | $k$ | $a_0$ | $t_b$ | $q$ | $t_2$ | $t'_2$ | dark | $J_\infty$ | $D_\infty P$ |
| --- | --- | --- | --- | --- | --- | --- | --- | --- | --- | --- | --- | --- | --- | --- |
| AFG | 34.66 | Mar 18 | Apr 3 | Jul 7 | Jun 28 | 0.995 | 68.9 | 36 | 0.16 | 4.0 | 24.8 | 7.1 | 1.00% | 1725 |
| ALB | 2.88 | Mar 11 | Mar 22 | Jul 17 | Jul 19 | 0.988 | 24.4 | 1 | 0.14 | 4.7 | 34.2 | 6.8 | 2.41% | 346 |
| AND | 0.08 | Mar 19 | Mar 25 | Mar 23 | Apr 4 | 0.931 | 7.6 | 0 | 0.24 | 2.6 | 12.0 | 11.8 | 13.50% | 52 |
| ARG | 43.85 | Mar 6 | Apr 4 | Jul 13 | Jul 19 | 0.987 | 21.8 | 12 | 0.20 | 4.8 | 24.1 | 4.6 | 2.63% | 5774 |
| ARM | 2.93 | Mar 22 | Jun 9 | Jul 1 | Jul 19 | 0.897 | 1.1 | 14 | 0.30 | 11.4 | 43.7 | 4.0 | 19.78% | 2893 |
| AUT | 8.75 | Mar 9 | Mar 11 | Mar 31 | Apr 5 | 0.992 | 149.8 | 19 | 0.05 | 1.1 | 23.4 | 7.2 | 1.66% | 725 |
| BEL | 11.35 | Mar 4 | Mar 28 | Apr 3 | Apr 9 | 0.911 | 6.1 | 7 | 0.23 | 2.5 | 12.3 | 30.8 | 17.30% | 9822 |
| BFA | 18.65 | Mar 14 | Mar 20 | Mar 19 | Apr 5 | 0.999 | 1754.5 | 0 | 0.25 | 2.8 | 10.9 | 10.1 | 0.06% | 53 |
| BGR | 7.13 | Mar 7 | Mar 31 | Jul 15 | Jul 19 | 0.974 | 10.1 | 28 | 0.12 | 5.3 | 47.3 | 7.8 | 5.10% | 1828 |
| BLR | 9.51 | Mar 25 | Mar 19 | Jun 25 | Jul 19 | 0.975 | 40.9 | 32 | 0.03 | 1.4 | 55.1 | 1.6 | 4.91% | 2344 |
| BOL | 10.89 | Mar 23 | Apr 12 | Jun 24 | Jul 3 | 0.968 | 7.9 | 29 | 0.31 | 5.5 | 18.4 | 8.7 | 6.30% | 3428 |
| BRA | 207.65 | Mar 11 | Mar 15 | May 27 | Jul 13 | 0.919 | 12.1 | 46 | 0.02 | 1.4 | 64.9 | 8.3 | 15.83% | 164365 |
| CAF | 4.60 | May 24 | May 27 | Jun 15 | Jun 14 | 0.999 | 123.9 | 11 | 1.87 | 9.3 | 5.0 | 2.6 | 0.24% | 55 |
| CHE | 8.37 | Mar 1 | Mar 22 | Mar 23 | Apr 3 | 0.976 | 22.0 | 5 | 0.25 | 2.6 | 10.9 | 11.8 | 4.70% | 1969 |
| CHL | 17.91 | Mar 16 | Jun 7 | Jul 9 | Jul 8 | 0.910 | 2.0 | 11 | 0.22 | 7.5 | 38.3 | 5.5 | 17.43% | 15610 |
| CHN | 1378.67 | Jan 15 | Feb 1 | Apr 9 | Feb 16 | 0.999 | 1544.6 | 9 | 0.09 | 2.6 | 28.1 | 10.9 | 0.07% | 4765 |
| COL | 48.65 | Mar 15 | Apr 4 | Jul 16 | Jul 19 | 0.899 | 4.0 | 7 | 0.19 | 3.3 | 20.1 | 8.4 | 19.48% | 47385 |
| CUB | 11.48 | Mar 19 | Mar 22 | Apr 16 | Apr 15 | 0.999 | 631.5 | 21 | 0.25 | 2.9 | 11.5 | 7.1 | 0.15% | 86 |
| CZE | 10.56 | Mar 17 | Mar 25 | Apr 1 | Apr 11 | 0.997 | 235.1 | 8 | 0.08 | 1.7 | 21.4 | 5.3 | 0.69% | 365 |
| DEU | 82.67 | Mar 2 | Mar 23 | Apr 8 | Apr 11 | 0.989 | 57.7 | 11 | 0.15 | 2.2 | 15.1 | 9.0 | 2.21% | 9146 |
| DNK | 5.73 | Mar 8 | Mar 23 | Mar 28 | Apr 8 | 0.989 | 52.6 | 6 | 0.16 | 2.4 | 15.3 | 9.2 | 2.15% | 615 |
| DOM | 10.65 | Mar 12 | Mar 23 | Jul 17 | Jul 19 | 0.747 | 2.6 | 16 | 0.04 | 1.9 | 76.4 | 4.0 | 45.91% | 24442 |
| DZA | 40.61 | Mar 6 | Mar 31 | Apr 2 | Jul 19 | 0.977 | 18.4 | 9 | 0.05 | 3.3 | 65.2 | 10.0 | 4.48% | 9105 |
| ECU | 16.39 | Mar 7 | Mar 7 | Apr 25 | Jul 8 | 0.935 | 15.5 | 40 | 0.01 | 1.3 | 119.5 | 14.9 | 12.72% | 10422 |
| EGY | 95.69 | Mar 6 | Apr 3 | Jun 8 | Jun 22 | 0.994 | 52.5 | 5 | 0.22 | 4.4 | 19.8 | 10.5 | 1.20% | 5724 |
| ESP | 46.44 | Feb 26 | Mar 17 | Mar 25 | Mar 31 | 0.937 | 12.4 | 6 | 0.14 | 1.7 | 13.4 | 21.9 | 12.27% | 28498 |
| ETH | 102.40 | Mar 29 | Mar 29 | Jul 19 | Jul 10 | 0.999 | 215 | 4 | 1.29 | 21 | 16.2 | 4.7 | 0.06% | 315 |
| FRA | 66.90 | Feb 19 | Mar 27 | Apr 2 | Apr 5 | 0.954 | 10.0 | 8 | 0.22 | 3.0 | 14.2 | 28.5 | 9.01% | 30142 |
| GAB | 1.98 | Apr 16 | Apr 30 | Jun 8 | Jun 6 | 0.997 | 64.6 | 0 | 0.39 | 7.6 | 19.4 | 1.6 | 0.56% | 56 |
| GBR | 65.64 | Feb 29 | Mar 26 | Apr 2 | Apr 19 | 0.925 | 8.9 | 12 | 0.09 | 2.0 | 26.1 | 30.9 | 14.54% | 47719 |
| GHA | 28.21 | Mar 16 | Mar 21 | Jun 12 | Jun 18 | 0.999 | 464.9 | 0 | 0.22 | 4.4 | 19.9 | 1.2 | 0.13% | 190 |
| GRC | 10.75 | Mar 7 | Mar 24 | Mar 27 | Apr 7 | 0.998 | 191.9 | 6 | 0.16 | 3.8 | 24.6 | 10.1 | 0.38% | 203 |
| GTM | 16.58 | Mar 28 | Jun 11 | Jul 7 | Jul 6 | 0.985 | 9.7 | 10 | 0.48 | 9.4 | 19.9 | 9.0 | 3.03% | 2513 |
| HND | 9.11 | Mar 22 | Mar 28 | Jun 24 | Jul 14 | 0.978 | 26.0 | 17 | 0.11 | 2.5 | 23.5 | 6.6 | 4.28% | 1952 |
| HRV | 4.17 | Mar 19 | Apr 11 | Apr 11 | Apr 28 | 0.996 | 72.3 | 17 | 0.04 | 4.8 | 122.5 | 6.3 | 0.79% | 165 |
| IND | 1324.17 | Mar 6 | Mar 21 | Jun 8 | Jun 28 | 0.997 | 189.9 | 36 | 0.12 | 2.4 | 20.8 | 5.9 | 0.61% | 40316 |
| IRL | 4.77 | Mar 7 | Mar 13 | Apr 17 | Apr 16 | 0.962 | 21.2 | 29 | 0.11 | 1.7 | 16.9 | 13.7 | 7.41% | 1768 |
| IRN | 80.28 | Feb 12 | Mar 8 | Jul 14 | Jul 19 | 0.819 | 3.1 | 17 | 0.04 | 2.3 | 84.0 | 11.5 | 33.82% | 135755 |

**Table III.** Continuation of Tab. II.

| country | $P/M$ | $t = 0$ | $t_a$ | $t_m$ | $t_0$ | $k$ | $a_0$ | $t_b$ | $q$ | $t_2$ | $t'_2$ | dark | $J_\infty$ | $D_\infty P$ |
| --- | --- | --- | --- | --- | --- | --- | --- | --- | --- | --- | --- | --- | --- | --- |
| ITA | 60.60 | Feb 15 | Mar 13 | Mar 20 | Apr 1 | 0.940 | 10.2 | 8 | 0.14 | 2.2 | 17.6 | 28.7 | 11.70% | 35442 |
| KOR | 25.37 | Feb 14 | Feb 16 | Mar 14 | Mar 21 | 0.999 | 989.6 | 13 | 0.07 | 1.2 | 17.4 | 4.3 | 0.22% | 285 |
| KWT | 4.05 | Apr 6 | May 5 | May 22 | Jun 6 | 0.986 | 21.8 | 20 | 0.09 | 4.5 | 51.0 | 1.5 | 2.83% | 573 |
| LBN | 6.01 | Mar 4 | Mar 31 | Jul 19 | Jul 19 | 0.999 | 95.5 | 2 | 0.09 | 9.9 | 105.3 | 3.6 | 0.29% | 88 |
| LUX | 0.58 | Mar 11 | Mar 19 | Apr 4 | Apr 4 | 0.981 | 29.5 | 7 | 0.21 | 2.4 | 12.1 | 4.0 | 3.82% | 111 |
| MAR | 35.28 | Mar 10 | Mar 24 | Mar 28 | Apr 17 | 0.999 | 628.8 | 13 | 0.03 | 2.5 | 82.8 | 3.6 | 0.18% | 316 |
| MDA | 3.55 | Mar 20 | Mar 25 | Jun 1 | Jun 15 | 0.973 | 25.9 | 23 | 0.06 | 1.9 | 33.7 | 7.0 | 5.40% | 960 |
| MEX | 127.54 | Mar 13 | Mar 24 | Jul 19 | Jun 24 | 0.954 | 16.2 | 42 | 0.06 | 1.8 | 31.1 | 25.4 | 8.99% | 57326 |
| MKD | 2.08 | Mar 16 | Mar 31 | Jun 25 | Jul 19 | 0.927 | 5.4 | 9 | 0.11 | 3.4 | 33.1 | 10.1 | 14.29% | 1488 |
| MRT | 4.30 | May 6 | Jun 1 | Jun 3 | Jun 7 | 0.996 | 66.4 | 4 | 0.29 | 5.1 | 17.9 | 5.3 | 0.82% | 175 |
| MYS | 31.19 | Mar 10 | Mar 17 | Mar 15 | Mar 30 | 0.999 | 2064.2 | 7 | 0.11 | 1.7 | 16.2 | 2.8 | 0.08% | 123 |
| NGA | 185.99 | Mar 23 | Apr 25 | Jun 9 | Jun 24 | 0.999 | 405.4 | 21 | 0.16 | 5.2 | 33.1 | 4.7 | 0.13% | 1214 |
| NLD | 17.02 | Mar 1 | Mar 22 | Mar 31 | Apr 8 | 0.963 | 15.4 | 5 | 0.20 | 2.4 | 12.6 | 23.7 | 7.26% | 6182 |
| NPL | 28.98 | May 10 | Jun 2 | May 29 | Jun 20 | 0.999 | 953.4 | 3 | 0.34 | 7.7 | 22.5 | 0.5 | 0.04% | 55 |
| PAK | 193.20 | Mar 11 | Apr 8 | Jun 11 | Jun 13 | 0.997 | 101.1 | 17 | 0.22 | 4.0 | 18.2 | 4.4 | 0.69% | 6677 |
| PAN | 4.03 | Mar 15 | Mar 26 | Jul 10 | Jul 19 | 0.848 | 3.5 | 10 | 0.09 | 2.5 | 35.5 | 4.8 | 28.80% | 5808 |
| PER | 31.78 | Mar 13 | Mar 6 | Jul 16 | Jul 19 | 0.703 | 3.2 | 47 | 0.03 | 1.3 | 68.9 | 10.1 | 52.66% | 83666 |
| PHL | 103.32 | Mar 5 | Mar 23 | Jul 5 | Jul 19 | 0.996 | 142.4 | 19 | 0.04 | 2.5 | 58.0 | 5.7 | 0.78% | 4024 |
| POL | 37.95 | Mar 6 | Apr 2 | Apr 17 | Jun 5 | 0.993 | 63.1 | 20 | 0.05 | 3.4 | 67.0 | 8.3 | 1.30% | 2474 |
| PRT | 10.33 | Mar 11 | Mar 14 | Mar 31 | Apr 22 | 0.982 | 86.4 | 16 | 0.02 | 0.9 | 38.8 | 7.1 | 3.67% | 1896 |
| ROU | 19.71 | Mar 15 | Mar 17 | Jul 14 | Jul 19 | 0.909 | 13.8 | 21 | 0.02 | 1.1 | 79.1 | 11.7 | 17.60% | 17343 |
| RUS | 144.34 | Mar 18 | Mar 18 | May 22 | Jul 5 | 0.984 | 60.7 | 44 | 0.02 | 1.4 | 61.8 | 3.4 | 3.12% | 22543 |
| SEN | 15.41 | Mar 28 | May 9 | Jun 8 | Jun 24 | 0.998 | 79.9 | 1 | 0.55 | 11.5 | 21.0 | 4.3 | 0.30% | 232 |
| SMR | 0.03 | Mar 3 | Mar 13 | Mar 13 | Mar 19 | 0.867 | 2.9 | 0 | 0.41 | 3.5 | 10.4 | 12.0 | 25.32% | 42 |
| SOM | 15.01 | Apr 6 | Apr 19 | Apr 15 | May 4 | 0.999 | 591.7 | 0 | 0.24 | 3.7 | 15.2 | 6.0 | 0.13% | 95 |
| SRB | 7.06 | Mar 15 | Mar 25 | Jul 19 | Jul 19 | 0.931 | 10.1 | 15 | 0.03 | 1.9 | 73.7 | 5.1 | 13.50% | 4763 |
| SWE | 9.90 | Mar 7 | Mar 26 | Apr 14 | May 1 | 0.935 | 10.1 | 16 | 0.07 | 2.1 | 33.6 | 14.7 | 12.75% | 6311 |
| TCD | 15.48 | Apr 21 | Apr 28 | Apr 30 | May 4 | 0.999 | 1387.0 | 3 | 0.22 | 2.1 | 9.2 | 16.9 | 0.10% | 75 |
| THA | 68.86 | Mar 17 | Mar 29 | Mar 26 | Apr 1 | 0.999 | 3463.0 | 0 | 0.56 | 4.8 | 8.6 | 3.6 | 0.02% | 58 |
| TUN | 11.40 | Mar 15 | Mar 30 | Mar 26 | Apr 1 | 0.999 | 659.7 | 6 | 0.29 | 4.7 | 16.4 | 7.3 | 0.09% | 50 |
| TUR | 79.51 | Mar 12 | Mar 17 | Apr 9 | May 6 | 0.990 | 144 | 21 | 0.01 | 1.0 | 108.3 | 5.1 | 1.97% | 7851 |
| USA | 323.13 | Feb 24 | Mar 23 | Apr 10 | May 16 | 0.938 | 10.2 | 23 | 0.03 | 2.1 | 81.6 | 7.8 | 12.18% | 196763 |
| ZAF | 55.91 | Mar 22 | May 18 | Jul 14 | Jul 19 | 0.968 | 6.5 | 21 | 0.32 | 6.6 | 21.1 | 3.7 | 6.34% | 17728 |

## Notes

### Competing Interest Statement

The authors have declared no competing interest.

### Funding Statement

This research did not receive any funding.

### Author Declarations

All ethical guidelines have been followed according to ETH Zurich regulations.

